# Tocilizumab efficacy in COVID-19 patients is associated with respiratory severity-based stages

**DOI:** 10.1101/2021.03.04.21252167

**Authors:** Melchor Alvarez-Mon, Angel Asunsolo, José Sanz, Benjamin Muñoz, José Alberto Arranz-Caso, Maria Novella Mena, Cristina Hernández-Gutiérrez, Jorge Navarro, Maria Cristina Lozano Duran, Juan Arévalo Serrano, Rocío Henche Sanchez, Lara Bravo Quiroga, Julio Flores Segovia, Marta Garcia Sanchez, Aida Gutierrez Garcia, Ana Pérez, Marta Herrero, Nieves Plana, Daniel Troncoso, Gorjana Rackov, Carlos Martinez-A, Dimitrios Balomenos, for The Medicine-COVID19 HUPA group

## Abstract

**Background:** Tocilizumab treatment is investigated, and effectiveness in ICU-admitted COVID-19 patients has been reported. Although controversy exists regarding the efficacy of tocilizumab treatment, it has been suggested that tocilizumab might show positive results depending on patient severity status. We examined an association between tocilizumab and distinct disease severity stages.

**Methods and Findings:** From March 3 to March 23 2020, 494 consecutively admitted COVID-19 patients received tocilizumab or standard treatment alone. Data were obtained retrospectively. Clinical respiratory severity (CRS) stages were defined by patient oxygenation status and were also associated to scores of WHO clinical progression scale. We categorized patients in three stages, mild/moderate CRS1 (FiSpO_2_<0.35; WHO score 5), moderate/severe CRS2 (FiO_2_=0.5/high flow mask; WHO score 6) and severe/critical CRS3 (FiO_2_<80%/high flow/prone position or mechanical ventilation; score>6). The primary outcome was the composite of death or ICU admission in patients of stages CRS1, CRS2, and CRS3, as well as in total patients. We also addressed mortality alone in total patients. Kaplan-Maier curves, Cox proportional regression and inverse probability weighting marginal structural models were used. We conducted the study from March 3 to April 7 2020 with broad-ranged severity patients; 167 tocilizumab-treated and 327 untreated. CRS1 patients showed no apparent benefit after treatment, while the risk of the primary outcome was greatly reduced in CRS2 treated participants ((HR=0.22; 95% CI (0.16-0.44)). Moreover, tocilizumab treatment was associated with significantly decreased CRS2 patient proportion that reached the outcome compared to non-treated controls (27.8.0% vs. 65.4%; p<0.001). Severe/critical CRS3 patients, also showed benefit after treatment (HR=0.38; 95% CI (0.16-90)), although not as robust as was that of CRS2 treated individuals. Tocilizumab was associated with reduced outcome risk in total patients (HR=0.42; 95% CI (0.26-0.66)) after CRS adjustment, but not if CRS classification was not accounted as confounding factor (HR=1.19; 95% CI (0.84-1.69)). The outcome of mortality alone upon tocilizumab treatment was significant (HR=0.58; 95% CI (0.35-0.96)) after accounting for CRS classification.

**Conclusions:** Tocilizumab treatment is associated with reduced COVID-19 escalation in CRS2 patients, suggesting efficacy in moderate/severe non-ICU-admitted patients. CRS classification could represent an essential confounding factor in evaluating tocilizumab in studies of broad-ranged severity patients.

## INTRODUCTION

Severe acute respiratory syndrome coronavirus-2 (SARS-CoV-2), has spread worldwide and is considered pandemic. Approximately 10-15% of patients, primarily older individuals (1, 2) (> 60 years of age) develop severe COVID-19 (Coronavirus disease 2019) pneumonia that requires hospitalization and/or intensive care unit (ICU) admission and suffer high mortality rates.

COVID-19 progression is related to cytokine storm linked hyperinflammation, driving acute respiratory distress syndrome (ARDS), multiple organ failure and sepsis [3]. Hyperinflammation and elevated cytokine and chemokine levels are associated with increased risk of death in patients with COVID-19 [4], providing rationale for immunomodulatory therapy [5,6]. Tocilizumab is a humanized monoclonal antibody that binds the soluble and membrane bound IL-6 receptor [7]. It has been approved for autoimmune disease treatment [8] and for CAR-T-therapy-related cytokine release syndrome [9,10].

Initial pilot studies suggested a possible tocilizumab effect in COVID-19 treatment [11]. More recent cohort studies showed effectiveness of tocilizumab in severe/critical patients [12–15], while clinical trials established no effect of tocilizumab in early stage patients (mild/moderate) [16–18]. Some of the trials with negative results have been criticized [19,20] and limitations to these studies have been emphasized, generating controversy about the effect of tocilizumab [21]. Preliminary data from the COVACTA clinical trial [22] raised important questions about the efficacy of tocilizumab in reducing the duration of hospital and ICU stay. However, the EMPACTA trial suggested tocilizumab efficacy in delaying disease escalation [23]. Recently, preliminary data from the REMAP-CAP trial reported benefit in the most severely ill patients [24], while the RECOVERY trial demonstrated that tocilizumab reduces mortality in patients receiving corticosteroids [25]. Recent meta-analysis found an association between tocilizumab and improved outcomes in moderate and severe patients (non-ICU), as well as in critically ill patients (ICU) [26].

We conducted a retrospective cohort study of 494 consecutively admitted COVID-19 patients that were classified in stages according to their Clinical Respiratory Severity (CRS). We provide evidence that tocilizumab is associated with reduced disease escalation in a CRS-depending manner, and that CRS classification is a critical confounding factor to be accounted for upon evaluation of tocilizumab’s therapeutic potential in the global patient population.

## METHODS

### Patients

Patients positive for SARS-Cov-2 from **March 3 to March 23 2020** who met the conditions of (1) respiratory rate ≥ 30 breaths/min, (2) SpO_2_ ≤ 94% while breathing ambient air, and (3) thorax X-ray with opacities, were included in the study, as defined by the Diagnosis and Treatment Protocol for Novel Coronavirus Pneumonia (6th interim edition) [27].

We obtained demographic data, as well as comorbidities (cancer, diabetes mellitus, cardiovascular disease, chronic kidney disease, among others) from electronic health records. We registered clinical symptoms, time of disease onset and laboratory and radiological data.

Standard treatment (ST) included lopinavir/ritonavir, hydroxychloroquine, IFN-β and azithromycin in case of secondary infections (Supplementary Table 1). IFN-β administration depended on the comorbidity extent and on age (> 65 years old). Two doses of IFN-β (at admission and at 48 h, discontinued then after due to apparent patient worsening) were given to 89 tocilizumab-treated patients and to 90 controls. At admission, patients received oxygen support though low-flow nasal cannula to maintain SpO_2_>90%. Patients with increased oxygen needs were switched to high-flow oxygen mask (Venturi mask up to 50% FiO_2_). Mechanical ventilation (MV) was provided only to ICU admitted patients. Considering the critical situation at the hospital, access to ICU was limited and patients that would otherwise be admitted to ICU were treated on the ward.

### Study plan

We carried out an open cohort study from **March 3 to April 7 2020**, including patients that received tocilizumab (Supplementary Table 1) in addition to ST or only ST.

Patient selection for tocilizumab treatment depended on the study design, the availability of tocilizumab and on ethical decisions, based on patient evolution during the time between admission and treatment. ST was given to all patients of both groups upon admission. Patients received tocilizumab after four-day observation (60% of patients at 4 day; for all patients mean ± SD, 4.06±0.099). Initial observation depended on SpO_2_ levels and clinical evaluation. Patients with rapidly increasing oxygen needs or radiological or clinical worsening were prioritized to receive tocilizumab.

The study was approved by the institutional review board of the University Hospital Prince of Asturias (HUPA0406/20). All patients provided informed consent before starting treatment.

### Study definitions

The study was designed to examine the effect of tocilizumab in patients of wide-ranged severity separated in groups. After data collection we classified patients according to oxygenation needs (FiO_2_) at the time of treatment and considering the WHO clinical progression scale [28], after its publication. We defined three clinical respiratory severity (CRS) stages; (1) CRS1; mild/moderate patients on low-flow oxygen supplementation FiO2≤0.35, WHO score 5, (2) CRS2; moderate/severe patients on high-flow oxygen supplementation FiO2=0.50, WHO score 6, (3) CRS3; severe/critical patients on high-flow oxygen supplementation FiO2=0.50, prone position, or on MV in ICU at the day of treatment, WHO score >6. CRS3 patients were considered critical not only because of the critical respiratory status but also due to the severity of CRS3 patients which showed much higher proportions of death or ICU admission compared to CRS2 individuals.

Initially, patient severity was considered on the basis of SpO_2_ measurements on ambient air and/or after low-flow oxygen supply during initial observation (24 hours or less). The groups were defined by patients with SpO_2_>90% (corresponding to CRS1), 80%<SpO_2_<90% (corresponding to CRS2), and SpO_2_<80% (corresponding to CRS3). By the end of the study we considered that we had enough patients in order to proceed to statistical analysis. This characterization was based on reports, showing that ARDS patients with oxygen supplementation and SpO_2_<90% show increased mortality, compared to patients with SpO_2_>90% [29]. Also, COVID patients with dyspnea (frequently manifested at SpO_2_<90%) show propensity for disease escalation [30]. Furthermore, there is an association of SpO_2_<80% patients with severe hypoxemia and increased risk of developing hypoxia [31– 33].

### Outcomes of the study

The primary outcome of the study was composite of death or admission to the ICU after tocilizumab treatment of CRS1, CRS2 and CRS3 stage patients, as well as of the total patient population.

In the secondary analysis death alone was conducted as endpoint, counting all in-hospital deaths of the patients. Deaths that occurred after ICU admission were available from hospital records and were also included in the analysis.

### Statistical analysis

Descriptive statistics were used to summarize the data; age and days between onset of symptoms and hospital admission were reported as means and standard deviations, and laboratory results are reported as medians and interquartile ranges (IQR). Categorical variables were summarized as counts and percentages. Associations between patient characteristics and tocilizumab treatment were evaluated; in the case of categorical variables, we used a Pearson’s chi-squared or Fisher exact test, and in the case of quantitative variables, Student’s t test, the Mann-Whitney’s U test, or Wilcoxon’s test, as appropriate. No imputation was made for missing data and no sample size calculations were performed.

We conducted a Cox proportional regression model with fixed covariates of patients from the day of treatment to the day of ICU admission or death for the different patient CRS stages adjusting for age, sex and IFN-β treatment. For total patients Cox analysis was also adjusted for CRS stages (CRS1, CRS2, CRS3). The start date of the follow-up was the day they received the medication. There were no differences in censorship for hospital discharge in both treated and untreated groups. We tested the proportional-hazards assumption on the basis of Schoenfeld residuals. This model may produce biased effect estimates when there are time-dependent confounders, which are affected by previous treatment or exposure. In this study, the CRS status is both a time-dependent confounder of the causal effect of the treatment on survival and is affected by past treatment [34,35]. We thus conducted a marginal structural model to estimate the causal effect of tocilizumab on the severe COVID-19 patients. We carried out an unweighted pooled logistic regression, treating each person-day as an observation (model 2). We fitted a pooled logistic regression weighted by the inverse-probability-of-treatment (IPWT). Each patient in the above logistic model received a time-varying weight inversely proportional to the estimated probability of having his/her own observed history of tocilizumab initiation, as described [34,36]. This weighting means that observations on the same subject will correlate, we therefore used the robust standard errors for clustering. Follow-up start day was the day of admission.

All models were adjusted for CRS stages (unless otherwise mentioned), sex, age, and IFN-β treatment. All statistical analyses were performed using STATA v14 SE or SPSSv26.

## RESULTS

We included 494 consecutively admitted patients, positive for SARS-CoV-2 and COVID-19 pneumonia. 167 patients received tocilizumab, while 327 did not (Figure 1). Age, gender distribution and frequency of comorbidities were similar for both groups (Table 1). Disease was classified in stages CRS1, CRS2 and CRS3 according to severity. Due to clinical selection, CRS2 and CRS3 stages had higher proportions of treated than untreated patients (Table 1). Occurrence of secondary infections was minimal (<5%) by both patient groups, and no opportunistic infections were recorded.

**Figure 1.**
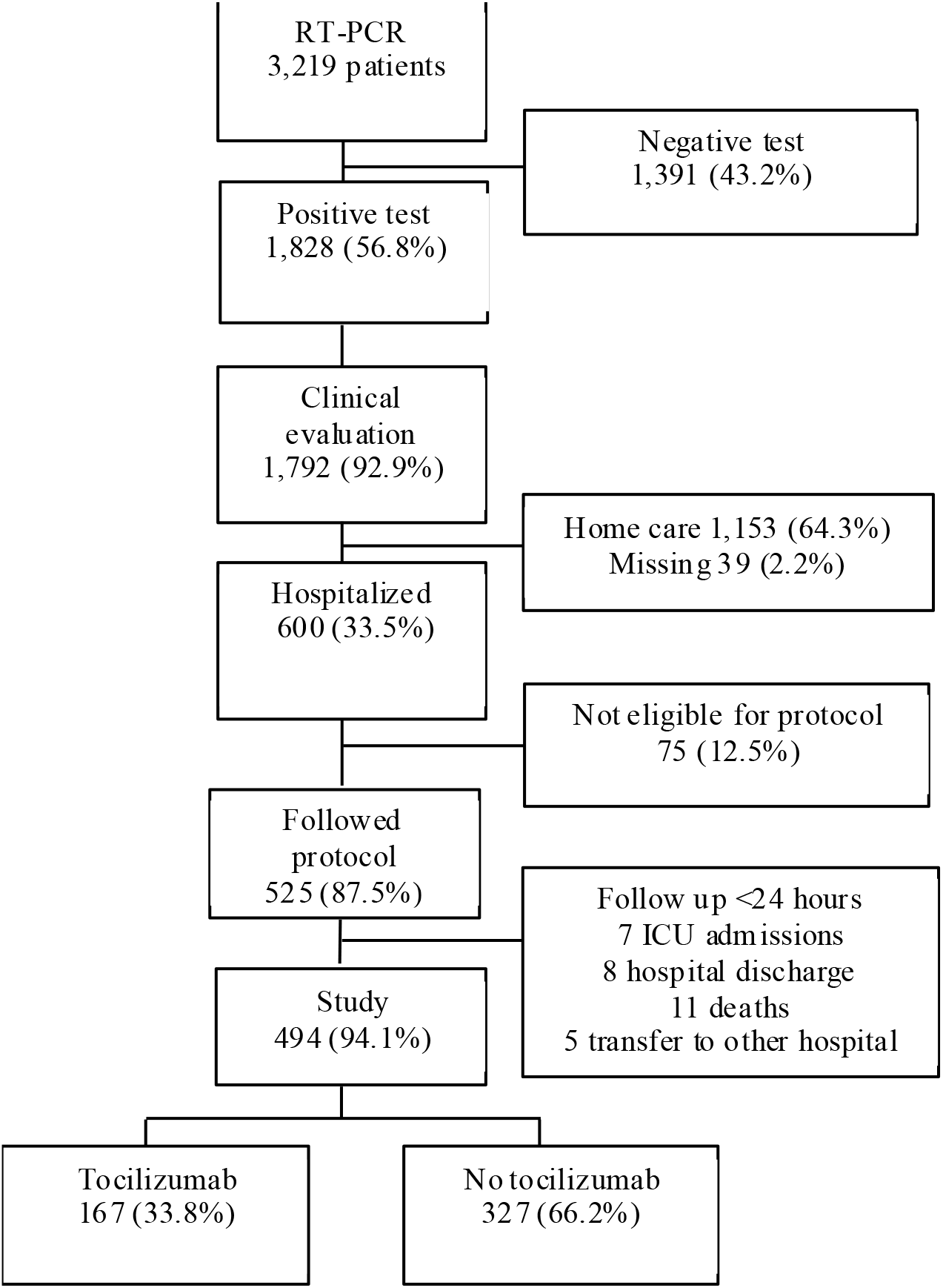
Selection of patient groups for tocilizumab treatment.

**Table 1.** Descriptive characteristics of the patient population

|  | Tocilizumab | No<br>tocilizumab | p |
| --- | --- | --- | --- |
| N (patients) | 167 | 327 |  |
| Age (years), mean (SD)* | 66.24<br>(11.38) | 66.93<br>(15.76) | 0.62 |
| Gender (male) % | 67.1 | 55 | 0.012 |
| Days between onset symptoms<br>and admission,<br>mean (SD)* | 7.29 (4.88) | 6.74 (5.62) | 0.262 |
| Onset symptoms (% of patients) |  |  |  |
| Fever | 91.2 | 85.3 | 0.078 |
| Cough | 77.4 | 78.1 | 0.86 |
| Throat pain | 5.8 | 7.7 | 0.46 |
| Dyspnea | 72.6 | 65.2 | 0.12 |
| Vomiting | 9.8 | 13 | 0.33 |
| Diarrhea | 28.7 | 24 | 0.30 |
| Comorbidities (% of patients) |  |  |  |
| Cancer | 6.5 | 10.5 | 0.17 |
| Diabetes | 27.4 | 23.8 | 0.42 |
| Cardiac or cerebrovascular disease | 55.7 | 56.8 | 0.83 |
| Chronic hepatitis | 3.8 | 3.7 | 0.95 |
| COPD | 5.2 | 5.7 | 0.81 |
| Chronic kidney disease | 7.1 | 9.8 | 0.34 |
| Neurological disease | 8.3 | 12.2 | 0.22 |
| Other treatments (% of patients) |  |  |  |
| Interferon | 53.3 | 27.5 | <0.001 |
| CRS (% of patients) |  |  | <0.001 |
| 1 | 26.9 | 88.6 |  |
| 2 | 53.9 | 8 |  |
| 3 | 19.2 | 3.4 |  |

Pearson's chi-squared test. \* Mann-Whitney U test

### CRS stage characteristics

CRS definition was validated after data collection. First, age, sex and comorbidities were found balanced within the CRS1, CRS2 and CRS3 stages (Table 2), indicating no bias in CRS definition. Second, CRP and HDL levels increased according to disease severity (CRS1<CRS2<CRS3), while lymphocyte concentrations were inversely correlated to this pattern (Table 2). These results support the CRS stage definition, since inflammation-associated factors are associated to COVID-19 severity [3], and hypoxemia could exacerbate inflammatory progression [37]. Third, SpO_2_ levels at baseline and after oxygen supplementation followed the CRS stage severity graduation (Table 2). SpO_2_/FiO_2_ ratios were obtained for CRS1, CRS2 and CRS3 patients on the basis of SpO_2_ limit values, as FiO_2_ was ≤35% for CRS1, while all CRS2 and CRS3 patients received FiO_2_=50% or MV after ICU admission (Table 2). CRS stages were correlated to ARDS development, since SpO_2_/FiO_2_ cutoffs of 100<200<300 predict ARDS [38]. Thus, for CRS1, SpO_2_/FiO_2_ reflected lower probability for ARDS and higher probability for CRS2 and CRS3 stages. Equivalent PaO_2_/FiO_2_ ratios were estimated from SPO_2_/FiO_2_ values [39] (Table 2) and were cautiously considered, since they were not directly obtained. Thus, CRS1 stage corresponded to mild, CRS2 to moderate and CRS3 to severe ARDS, according to established criteria [40,41]. Based on the above results, we considered the disease of CRS1 as mild/moderate, CRS2 as moderate/severe and CRS3 as severe/critical, since at baseline CRS3 patients received high-flow oxygen or MV after ICU admission.

**Table 2.**
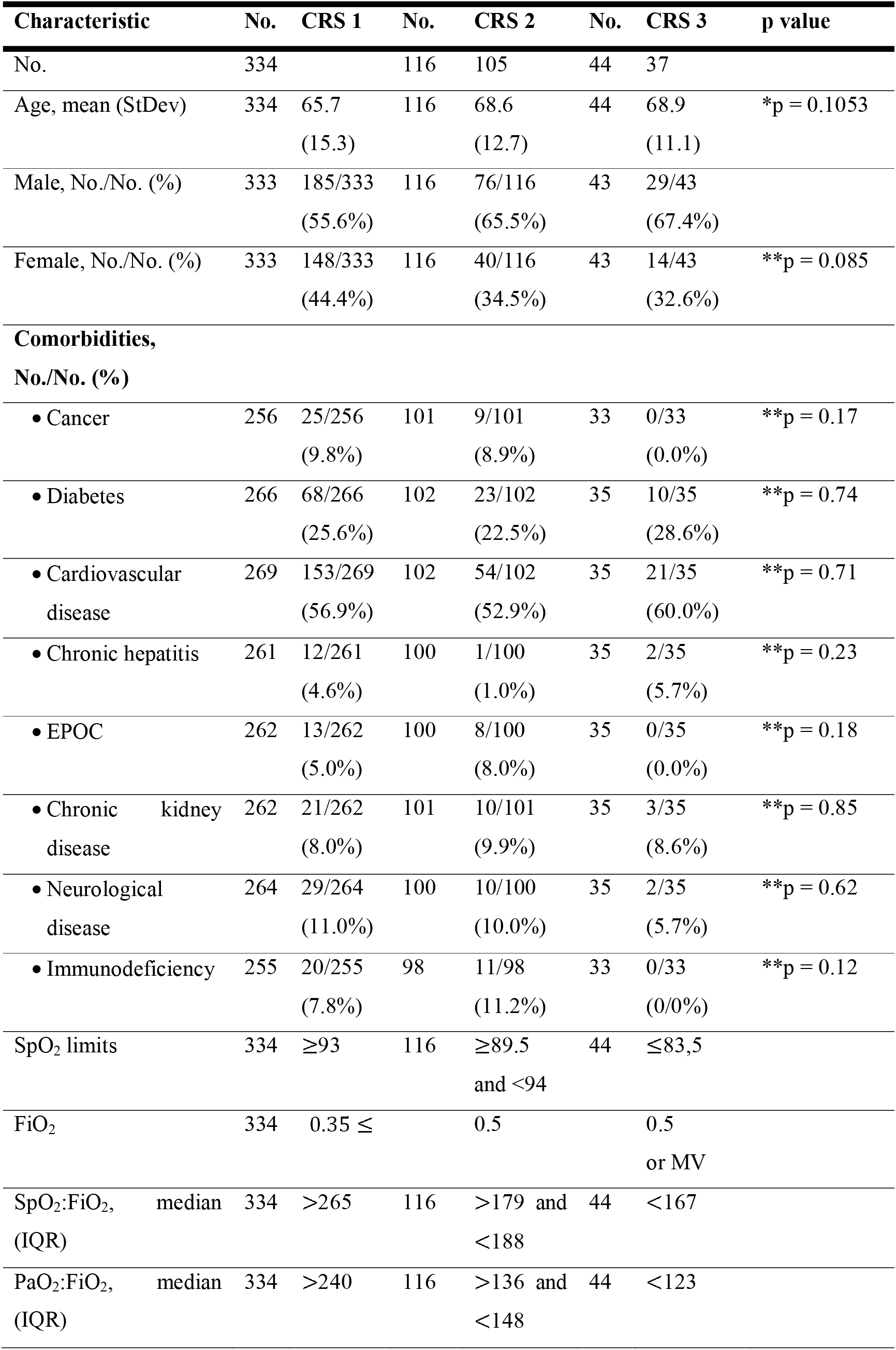
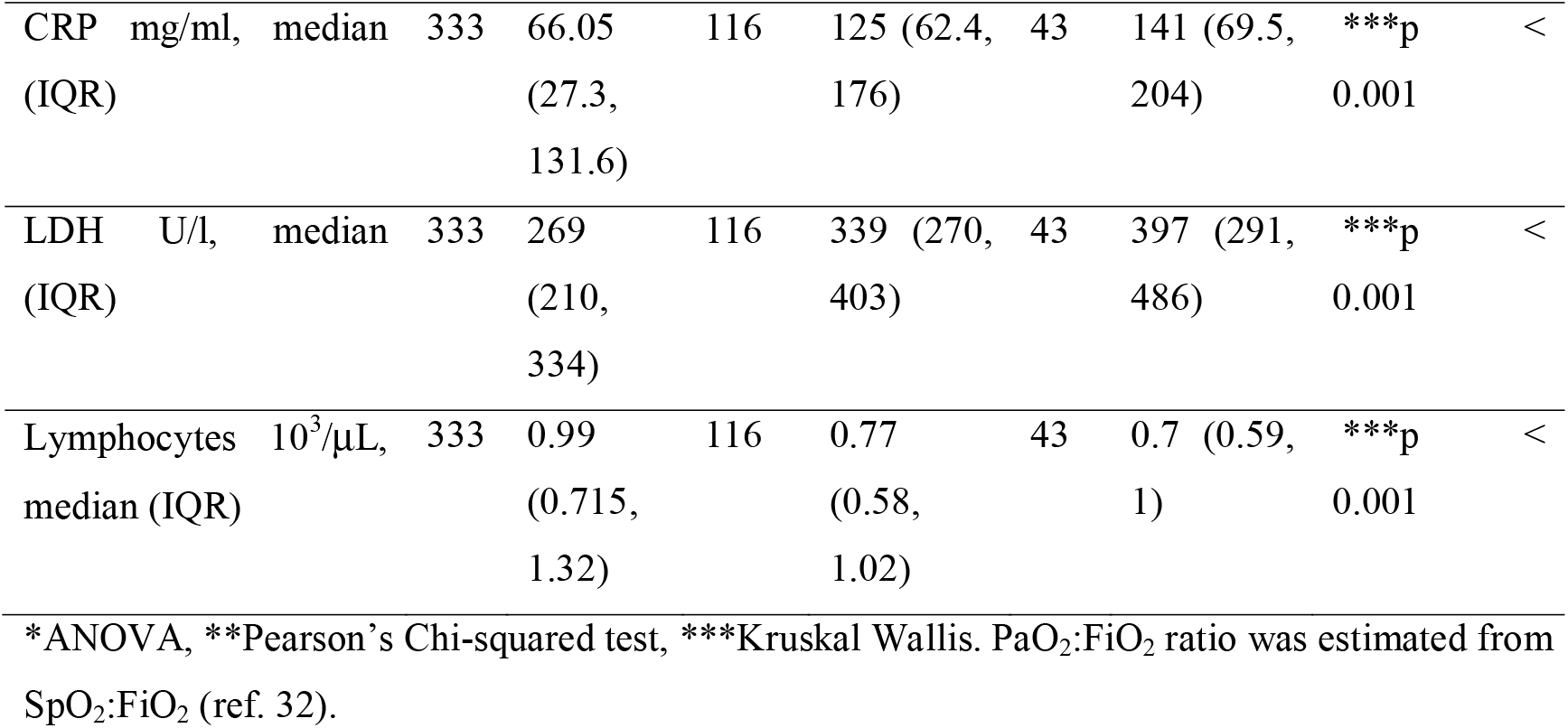
Baseline patient characteristics of CRS stages.

### Outcomes

Analysis of the tocilizumab effect on the primary outcome for CRS1, CRS2 and CRS3 patients included Kaplan-Meier survival curves and multivariate Cox regression after adjusting for age, sex and the IFN-β impact. In CRS1 patients, tocilizumab did not show an apparent effect in lowering the outcome risk (HR=0.76 95% CI (0.34, 1.70)) (Figure 2A). Although the percentage of CRS1 patients that reached the endpoint was reduced (23.5% untreated *vs*. 15.6% treated), the results lacked significance (Figure 2B). On the other hand, the data show significantly delayed outcome in tocilizumab treated CRS2 and CRS3 patients (Figure 2A). A benefit was probable to occur for CRS2 treated patients (HR=0.22 95% CI (0.16, 0.44)) and for CRS3 patients (HR=0.38 95% CI (0.16, 0.90)) (Figure 2A). Moreover, our data indicated that tocilizumab could significantly reduce the proportion of CRS2 patients that reached the endpoint (65.4% untreated vs. treated 27.8% patients; p<0.001) (Figure 2B). Such reduction appeared to be not significant for treated CRS3 patients, possibly due to the low number of untreated patients in CRS3. These data suggest that tocilizumab could be most effective when administered to CRS2 stage patients.

**Figure 2.**
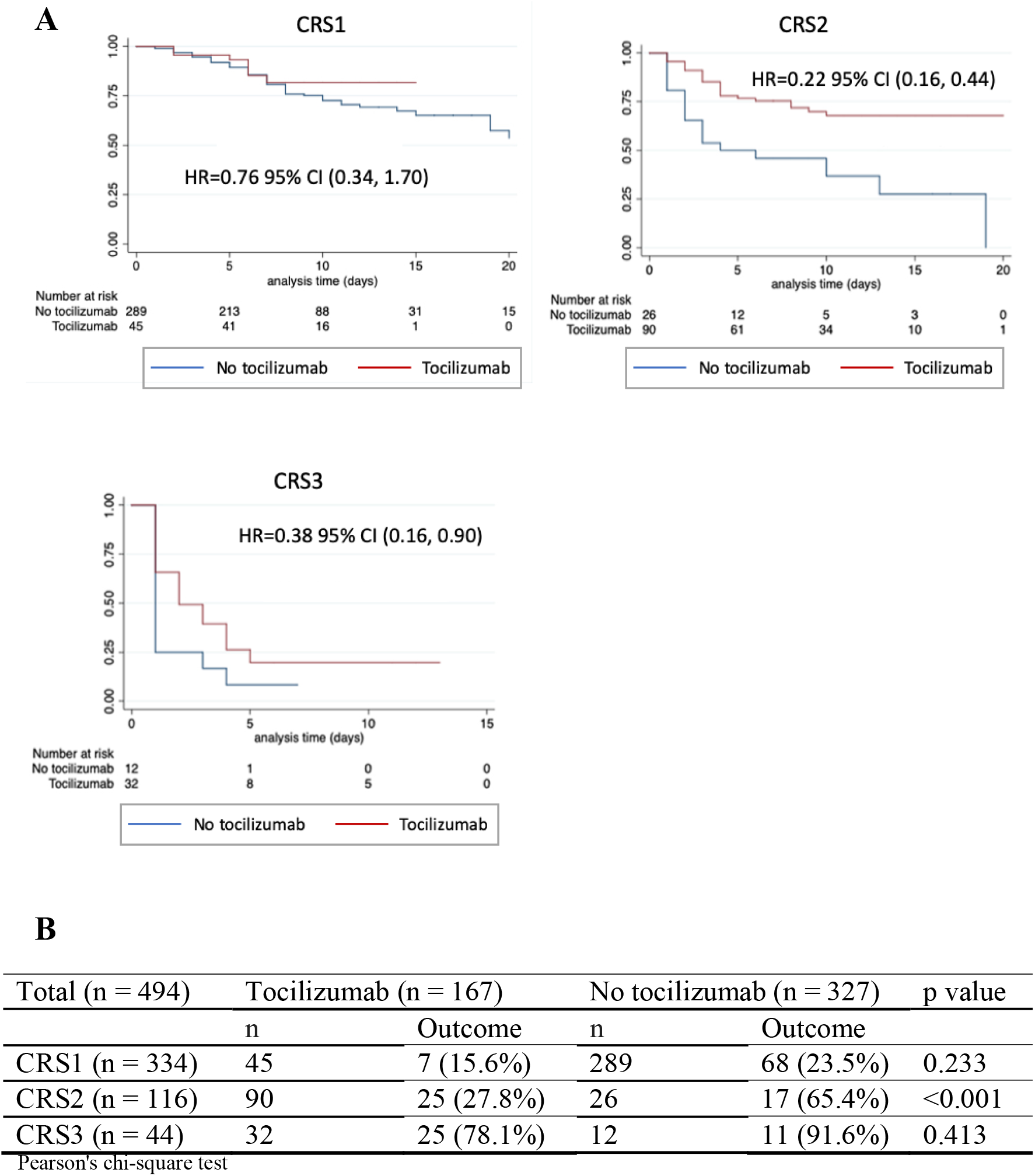
**(A)** Kaplan-Meier estimation of clinical improvement in CRS1, CRS2 and CRS3 patients. Multivariate Cox regression analysis; p = 0.334 (CRS1); p< 0.001 (CRS2); p = 0.018 (CRS3). **(B)** Numbers and proportions of tocilizumab treated and untreated patients that reached the outcome; Pearson’s chi-square test.

In the entire treated population, tocilizumab greatly reduced CRP levels and elevated lymphocyte concentrations 48 h post-treatment (Supplementary Table 2), corroborating previous reports [3]. To evaluate the effect of tocilizumab in all patients with respect to the composite endpoint, we conducted a multivariate Cox proportional-hazards analysis and accounted for CRS stage classification as confounding variable, in addition to age, sex and the impact of IFN-β. We showed an association between tocilizumab treatment and reduced risk of reaching the composite outcome (Figure 3A and Table 3, model 1).

**Figure 3.**
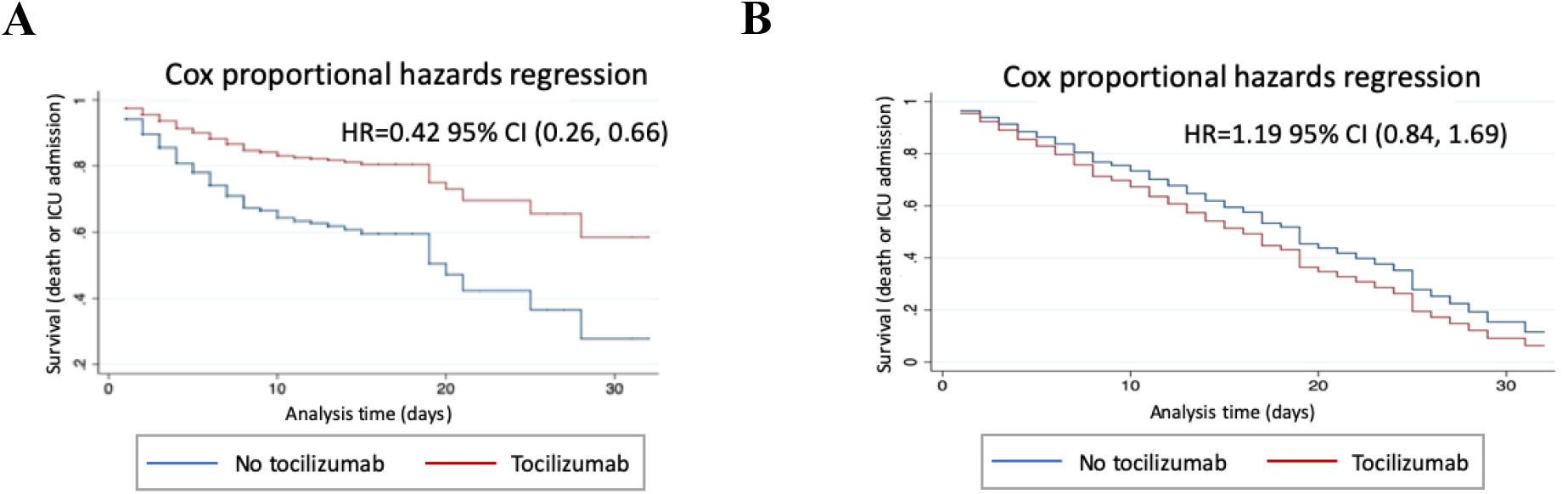
Increased probability of survival after tocilizumab treatment. Survival corresponds to improvement of the end event defined as death or ICU admission. HR values correspond to multivariate Cox regression analysis. **(A)** Adjusted for age, sex, comorbidities, other treatments and CRS stages, as in Table 3 (model 1); **(B)** Adjusted as in (A) but not for CRS stages.

**Table 3.** Multivariate regression models to estimate the effect of tocilizumab adjusted for the CRS stages.

|  | **Model 1** |  |  |  | **Model 2** |  |  |  | **Model 3** |  |  |  |
| --- | --- | --- | --- | --- | --- | --- | --- | --- | --- | --- | --- | --- |
|  | HR | [95% CI] |  | p | HR | [95% CI] |  | P | HR | [95% CI] |  | p |
| Tocilizumab | **0.42** | **0.26** | **0.66** | <0.001 | **0.22** | **0.09** | **0.59** | 0.002 | **0.17** | **0.06** | **0.47** | <0.001 |
| Age (years) | 1.03 | 1.01 | 1.04 | <0.001 | 1.02 | 1.00 | 1.04 | 0.023 | 1.03 | 1.00 | 1.05 | 0.020 |
| Sex (male) | 1.26 | 0.90 | 1.76 | 0.183 | 1.19 | 0.75 | 1.91 | 0.46 | 1.53 | 0.91 | 2.59 | 0.107 |
| CRS | - |  |  |  | - |  |  |  | - |  |  |  |
| 2 | 2.80 | 1.75 | 4.47 | <0.001 | 8.45 | 4.40 | 16.22 | <0.001 | 10.42 | 5.23 | 20.76 | <0.001 |
| 3 | 12.75 | 7.65 | 21.23 | <0.001 | 23.14 | 12.83 | 41.74 | <0.001 | 27.23 | 1.47 | 64.66 | <0.001 |
| Other treatments: |  |  |  |  |  |  |  |  |  |  |  |  |
| Interferon | 1.87 | 1.33 | 2.62 | 0.001 | 1.21 | 0.75 | 1.95 | 0.428 | 1.55 | 0.91 | 2.64 | 0.108 |

Model 1 Cox proportional regression
Model 2 Unweighted pooled logistic regression
Model 3 Marginal structural model. Pooled logistic regression weighted by IPWT

In secondary analysis, Cox evaluation including the above adjustments, also indicated reduced risk for mortality alone after tocilizumab treatment, considering all hospital deaths (HR=0.58 95% CI (0.35, 0.96)).

Disease status of COVID-19 patients was a decisive confounding factor to be accounted for regression analysis for our broad-ranged population. Notably, Cox analysis, that did not account for CRS stages but included all other mentioned confounding parameters, showed no effect of tocilizumab in reducing the risk for the composite outcome (HR=1.19 CI 95% (0.84-1.69)) (Figure 3B and Supplementary Table 3).

In addition to Cox based evaluation in all patients, we conducted an unweighted pooled logistic regression analysis for death or ICU admission and a weighted pooled logistic regression by IPWT (Table 3, models 2, 3). These models indicated that tocilizumab relates with a reduced risk in patient mortality or ICU admission, and that age, but not sex, significantly increased the risk for the end event. The risk for the outcome was significantly increased by IFN-β treatment, at least by model 1. Importantly, the three models confirmed that the patients’ CRS stage is a significant risk factor for death or ICU admission and confirmed that the CRS2 that CRS3 patients, as compared to CRS1, present a significantly higher probability of reaching the end event.

## DISCUSSION

In this retrospective cohort study we tested tocilizumab efficacy in COVID-19 patients according to their clinical respiratory severity, and we concluded that: (1) tocilizumab had no apparent effect in reducing the risk of composite outcome (death or ICU admission) mild/moderate CRS1 patients; (2) the benefit from tocilizumab treatment was particularly pronounced in patients categorized as CRS2 (moderate/severe) and to a lesser extent in CRS3 patients (severe/critical); (3) analysis of total patients as a single population required CRS stage adjustment to show beneficial effect for tocilizumab.

Measuring SpO_2_ is not invasive and was easier to perform under the critical conditions of the COVID-19 pandemic. SpO_2_-based CRS stage classification is reinforced by another study showing that after oxygen supplementation, SpO_2_>90% predicted survival, while SpO_2_<90% levels were associated with COVID-19 escalation [42]. Results obtained during the study, such as CRP levels, SpO_2_/FiO_2_ and estimated PaO_2_/FiO_2_ ratios further validated CRS stage classification and corroborated the stage severity CRS1< CRS2< CRS3. Further refining of CRS stages using additional oxygenation, inflammatory and clinical parameters might aid in optimizing prediction of the earliest point of tocilizumab delivery.

Current views on tocilizumab use suggest no effect on early stage hospitalized patients but therapeutic benefit in severe/critical patients [43–45]. We found that tocilizumab did not provide significant benefit on disease progression for mild/moderate CRS1 patients (estimated PaO_2_/FiO_2_>240 mm Hg). This concurs with randomized trials, which showed no benefit after tocilizumab treatment in patients with mild/moderate disease severity (200<PaO_2_/FiO_2_<300 mm Hg) [16–18].

For CRS2 patients, multivariate Cox analysis showed an association between tocilizumab treatment and reduced outcome (death or ICU) risk. The clearest risk reduction was detected for CRS2 stage treated-patients (estimated PaO_2_/FiO_2_<150 mm Hg), and this was further confirmed by significantly decreased proportion of patients that reached the endpoint. These results concurred with another study showing reduced risk of disease escalation in treated-patients with PaO_2_/FiO_2_<150 mm Hg [46].

Our results suggested a greater tocilizumab effect in CRS2 compared to CRS3 stage patients, supporting the view that tocilizumab increases effectiveness when delivered at earlier disease stages [45,47]. This might be explained by more severe respiratory deficiency in CRS3 compared to CRS2 patients. Effectiveness of tocilizumab in CRS3 stage is corroborated by other studies focused on severe/critical patients, showing beneficial effects of tocilizumab for patients on MV [48], or tocilizumab delivery two days after ICU admission [45]. Finally, as CRS2 patients did not require ICU admission at baseline, tocilizumab treatment could be an option for moderate/severe patients on high-flow oxygen supply.

When global patient population was considered, three models of regression analysis showed that tocilizumab was associated with a reduced risk of the composite outcome, after accounting for CRS stage classification as a confounding factor together with patient age, sex and other treatments. In addition, multivariate Cox regression analysis showed significantly lowered risk in the case that mortality alone was the outcome.

Adjustment for CRS stages was decisive for the estimated therapeutic effect of tocilizumab in all patients, since after excluding the CRS stage classification, as confounding variable, tocilizumab benefit was no longer evidenced.

Preliminary data from clinical trials COVACTA and EMPACTA (19-20) that included broad ranged patient populations, showed that tocilizumab was not as efficacious as in observational studies that focused on advanced disease stages. The COVACTA study did not meet the primary outcome, as tocilizumab did not reduce clinical worsening or mortality. The wide-ranged severity of the patients could be a drawback, considering that the inefficacy of tocilizumab in mild/moderate severity patients might have obscured relevant therapeutic effects [44]. The EMPACTA trial [50] showed reduced risk of MV or death, but not of mortality alone in tocilizumab-treated patients.

On the basis of our findings, we believe that future clinical studies could draw more accurate conclusions by considering disease stage classification and including it as a confounding factor in regression analyses.

Overall, using a single tocilizumab appears to be safe for the patients and does not increase secondary infections, as shown by us and other studies [17,45,50]. Perhaps a second consecutive tocilizumab delivery might result in increased secondary infections [12].

Our study has the following limitations. First, the study was performed in one hospital and was not randomized as tocilizumab treatment was prioritized for worsening patients within the same CRS stage. Second, CRS3 stage patient population did not get the support of MV due to the near collapse conditions in Spain at the time of the study. Third, our study was not blinded and therefore a bias for ICU admission could favor untreated patients; this limitation was minimized as analysis of overall hospital mortality as an endpoint showed benefit after tocilizumab treatment.

The strengths of this study include the newly applied patient classification in CRS stages. This later point is crucial since previous studies mostly found no tocilizumab effect when considering patients of wide range disease severity. Several analyses were performed, with consistent results across the models. Oxygen supplementation was uniform especially for the CRS2 stage, as the vast majority of the patients were managed on the floor (*i*.*e*. not in the ICU) on steady FiO_2_, avoiding the bias of oxygen supplementation in influencing the outcome. Finally, although differences in tocilizumab efficacy have been suggested to depend on disease stage, here we show this in a single study that encompassed three disease stages.

## Conclusions

Tocilizumab shows variable efficacy, depending on three COVID-19 clinical respiratory severity (CRS) stages. Treatment had no apparent benefit in CRS1 patients, but was associated with a remarkable benefit in CRS2 patients, suggesting that tocilizumab could target stages that did not require ICU care. These findings could contribute to define the optimal point for treatment initiation. In general terms, our results indicate that the CRS-associated efficacy of tocilizumab could eventually hinder evaluation of its therapeutic effect, and suggest the possibility to consider CRS patient classification as a confounding factor in analyzing the efficacy of tocilizumab. These conclusions could assist in designing and/or interpreting randomized clinical trials.

## Data Availability

All data generated and analyzed during this study are included in this article.

## Acknowledgements

We thank Catherine Mark for editorial assistance. This work was partially supported by grants from the Fondo de Investigación de la Seguridad Social-Instituto de Salud Carlos III (PI18/01726), Spain. Programa de Actividades de I+D de la Comunidad de Madrid en Biomedicina (B2017/BMD-3804), Madrid, Spain. Instituto de Salud Carlos III CIBER Enfermedades hepáticas y Digestivas, Spain. Agencia Estatal de Investigación. SAF2016-80803-R. Medicina individualizada traslacional, en inflamación y cáncer. Comunidad de Madrid. B2017/BMD-3804, Spain (to: MAM, CMA and DB). GR holds Juan de la Cierva grant (Ministerio de Ciencia e Innovación).

