## Supplemental Info for "Tocilizumab efficacy in COVID-19 patients is associated with respiratory severity-based stages"

### Supplement

### eMethods

#### eProtocols 1. Management protocol of tocilizumab treatment for patients with confirmed COVID 19 pneumonia at the University Hospital Prince of Asturias

| | |
|---|---|
| **General Information** | Therapeutic regimen adapted to the our clinical setup for Tocilizumab treatment (March 03, 2020)*<br>Adopted from the recommendations of the Ministry of Health, Government of Spain.<br>(*This protocol was in force during the data collection from patients included in the study, it has been periodically updated subsequently) |
| **Standard treatment** | Positive PCR patients with opacities on chest radiography, oxygen saturation <94% and respiratory rate ≥30 breaths/min were given standard treatment upon admission.<br>**Lopinavir / ritonavir** (kaletra® comp, 200/50 mg), 2 comp/12h, plus **Hydroxychloroquine** (dolquine® 200 mg tablets), loading dose: 2 comp/12h during the first day and followed by 1 comp/12h po], depending on availability, plus **Acethylcisteine** (flumil® ampoules 300 mg), 600 mg/12h. Additionally, **Beta Interferon 1B** (Betaferon® or Extavia® 250 mcg) subcutaneous/48h for patients with age >65 years and comorbidities.<br>Treatment duration according to clinical evolution** |
| **Tocilizumab treatment** | Patients as in standard treatment, with rapidly increasing O2 needs or change from low-flow nasal cannula to face mask or with clinical/radiological worsening.<br>Standard treatment plus **Tocilizumab** (Roactemra®Vial 200 mg / 10 ml), Single dose 200-800 mg (according to weight) in 100 ml SSF, 1 infusion in 1h. Active bacterial infection was excluded. |
| **Adverse effects protocol** | In case of digestive intolerance or lacking of lopinavir/ritonavir (Kaletra®), darunavir/cobicistat (Rezolsta® pills 800/150 mg) 1 pill/day, will be used.<br>In case of requiring administration through nasogastric probe: Lopinavir/ritonavir oral solution (80/20 mg/ml) 5 ml/12h will be administered. It must be a PVC probe<br>Before using these drugs, the existence of contraindications should be excluded according to the technical data sheet.<br>All patients must have a close follow-up of hematological, biochemical and electrocardiographic tests |
| **Updates** | **Acethylcisteine, although included in the protocol, was not used as standard treatment. Beta Interferon treatment was discontinued after second dose due to apparent negative results. |

### eTables

#### eTable 1. Diagnosis and treatment

| | | |
|---|---|---|
| **Diagnosis** | | |
| VIASURE SARS-CoV-2 Real Time PCR Detection Kit (nasopharyngeal swabs) | CerTest Biotec, Spain | |
| **Standard treatment** | | |
| Lopinavir/ritonavir | Kaletra®, Abbott, USA | 200/50 mg, 2 tablets/12h |
| hydroxychloroquine | Dolquine®, Laboratorios Rubio, Spain | 200 mg loading dose, 2 tablets/12h for the first day, and 1 tablet/12h PO, $8x10^6$ iu/48 hours IV |
| IFN-beta 1b | Betaferon, Bayer, Germany | Two doses; $8x10^6$ iu/48 hours IV |
| Azythromycin | Pipeline Pharma, Spain | 500 mg/24h IV for 5-8 days |
| Tocilizumab | Roactemra®, Roche, Switzerland | 200 mg/10 ml ampoules as single dose 400 mg for <60 kg; 600 mg for 60-80 kg; 800 mg for >80 kg; intravenously in 100 ml of 0.9% saline solution in 1 h |

eTable 2. Laboratory results of tocilizumab-treated and untreated patients

| | **Tocilizumab** | | No tocilizumab | | | **Tocilizumab** | | No tocilizumab | |
|---|---|---|---|---|---|---|---|---|---|
| | | **Administration day** | | **Administration day** | **p*** | **48 hours later** | **p**** | **48 hours later** | **p**** |
| | n | median (IQR) | n | median (IQR) | | median (IQR) | | median (IQR) | |
| CRP | 164 | 130.75 (76.9, 176.65) | 247 | 61.8 (21.9, 129.3) | <0.001 | 29.8 (13.7, 73.25) | <0.001 | 42 (14.5, 89.1) | <0.001 |
| LDH | 119 | 334 (279, 412) | 200 | 263 (204.5, 334.5) | <0.001 | 365 (272.5, 473) | 0.02 | 255 (208.5, 320.5) | 0.047 |
| Lymphocytes ($10^3$/ µL) | 166 | 0.78 (0.6, 1.02) | 251 | 1 (0.71, 1.36) | <0.001 | 0.8 (0.6, 1.16) | 0.041 | 1.1 (0.74, 1.49) | <0.001 |

*Mann-Whitney U test (between groups, tocilizumab-no tocilizumab), ** Wilcoxon signed-rank test (intra-group, before-after)

eTable 3. Cox proportional regression model estimating the effect of tocilizumab without adjusting for CRS stages

| **Model 1 (without CRS)** | HR | [95% CI] | | p |
|---|---|---|---|---|
| Tocilizumab | 1.19 | 0.84 | 1.69 | 0.333 |
| Age (years) | 1.03 | 1.02 | 1.04 | <0.001 |
| Sex (male) | 1.23 | 0.89 | 1.71 | 0.227 |
| Other treatments:<br>Interferon | 1.77 | 1.26 | 2.47 | 0.001 |

### Authors

Investigators of The Medicine-Covid19 HUPA group:

Ana Maria Culebras Lopez

Ana Maria Caro Leiro

Enrique Saiz Hervas

Jose Miguel Rodriguez Gonzalez

Elena Rabadan
